# Estimating dates of origin and end of COVID-19 epidemics

**DOI:** 10.1101/2021.01.19.21250080

**Authors:** Thomas Beneteau, Baptiste Elie, Mircea T. Sofonea, Samuel Alizon

**Author notes:** This article has been peer-reviewed and recommended by *Peer Community In Mathematical and Computational Biology* (https://doi.org/10.24072/pci.mcb.100004).

## Abstract

Estimating the date at which an epidemic started in a country and the date at which it can end depending on interventions intensity are important to guide public health responses. Both are potentially shaped by similar factors including stochasticity (due to small population sizes), superspreading events, and ‘memory effects’ (the fact that the occurrence of some events, e.g. recovering from an infection, depend on the past, e.g. the number of days since the infection). Focusing on COVID-19 epidemics, we develop and analyse mathematical models to explore how these three factors may affect early and final epidemic dynamics. Regarding the date of origin, we find limited effects on the mean estimates, but strong effects on their variances. Regarding the date of extinction following lockdown onset, mean values decrease with stochasticity or with the presence of superspreading events. These results underline the importance of accounting for heterogeneity in infection history and transmission patterns to accurately capture early and late epidemic dynamics.

## 1 Introduction

The ability to make robust epidemiological inferences or predictions strongly relies on the law of large numbers, which buffers the variability associated with individual processes. Many models of infectious diseases spread are deterministic and therefore assume that the number of infected hosts is large and above what has been termed the ‘outbreak threshold’ (Hartfield and Alizon, 2013). This assumption is violated at the beginning and end of an epidemic, where stochasticity may have a strong effect (Britton and Scalia Tomba, 2019).

In this study, we tackle two issues. First, we wish to estimate the date of origin of an epidemic in a country, focusing on the case of COVID-19 outside China. This question is important because the infection being imported, some cases may be detected before the reported beginning of an epidemic wave, which is somehow counter-intuitive to an audience not familiar with stochasticity. Furthermore, transmission often takes place before an epidemic wave is detected, as shown in several places using SARS-CoV-2 genomic data, e.g. Washington state in the USA (Bedford et al., 2020) or France (Danesh, Elie, et al., 2020). Second, we investigate how many days strict control measures need to last to ensure that the prevalence falls below key thresholds. Despite its public health implications, this latter question has rarely been investigated. There are some exceptions, for instance in the context of poliomyelitis (Eichner and Dietz, 1996), Ebola virus disease (Thompson et al., 2019), and MERS (Nishiura, Miyamatsu, et al., 2016) epidemics. However, these estimates neglect superspreading events and/or do not include non-Markovian effects (i.e. memory effects). Indeed, they often rely on ordinary differential equations, meaning that the probability of an event to occur (e.g. recovering from an infection) does not depend at all on the past (e.g. the number of days since the infection started). Recently, however, it has been shown that incorporating secondary cases heterogeneity can significantly lower the delay until an Ebola virus disease outbreak can be considered to be over (Djaafara et al., 2020).

Maintaining the lockdown so as to reach ‘zero-COVID’ requires extended effort because the incidence might oscillate at a low value due to stochasticity for a long period. However, in practice, and as illustrated by several countries, lockdown measures could be eased after the epidemic reaches a sufficiently low incidence. Indeed, when the number of cases is low enough, stricter contact tracing, as well as local control measures can be sufficient to stop the virus spread. For instance, in Taïwan or South Korea, the epidemic was controlled for months as long as the incidence was kept below 20 new cases per day (Max Roser and Hasell, 2020). In New Zealand, control measures were lifted only when the incidence reached 2 cases per day. This is why we investigate the time for incidence to reach given thresholds that can be greater than 0.

The COVID-19 pandemic led to an unprecedented publication rate of mathematical models, several of which involve stochasticity. For instance, Hellewell et al., 2020 analysed the initial steps of the outbreak to estimate the fraction of the transmission chains that had to be tracked to control the epidemics. Their results depend on the value of the basic reproduction number (denoted *R*_0_), which corresponds to the mean number of secondary infections caused by an infected individual in an otherwise fully susceptible population (Anderson and May, 1991), but also on individual heterogeneity. Indeed, if few individuals tend to cause a large number of secondary infections while the majority tends to cause none, the probability of outbreak emergence is much lower than if all individuals cause the same number of secondary infections (Lloyd-Smith et al., 2005). Accounting for this property, a study used the early COVID-19 outbreaks incidence data in different countries to estimate the dispersion of the distribution of individual *R*_0_ (Endo et al., 2020). Finally, Althouse et al., 2020 have also used stochastic modelling to explore the role of super-spreading events in the pandemic and its consequences on control measures.

Here, we develop an original discrete stochastic (DS) model, which features some of the known characteristics of the COVID-19 epidemics. The model is non-Markovian, which means that individual histories matter for the dynamics. More specifically, the probability that an event occurs (e.g. infecting another host) depends on the number of days spent in a state (e.g. being infected). Furthermore, following earlier studies (Hellewell et al., 2020), we account for the fact that not all hosts transmit on the same day post-infection. This is captured by assuming a distribution for the generation time, which is the time between infection dates of an ‘infector’ and an infected person. Since the time of infection is complicated to estimate, we approximate the generation time by the serial interval, which is the time between the onset of the symptoms in the ‘infector’ and that in the infected person (He et al., 2020; Nishiura, Linton, et al., 2020b). We also allow for heterogeneity in transmission patterns by assuming a negative binomial distribution of the secondary cases. To investigate the importance of stochasticity, we had to use deterministic models in addition to ours. To have memory effects in a deterministic setting, we reanalysed an earlier non-Markovian model (Sofonea et al., 2020) by setting the date of origin of the epidemic as the main free parameter. Finally, to remove both memory effects and stochasticity, we analyse a classical deterministic Markovian model, which is commonly used to analyse COVID-19 epidemics (Grant, 2020).

By comparing the outputs of these models, we explore the importance of stochasticity, individual heterogeneity, and non-Markovian effects on the estimates of the dates of origin and end of a nation-wide COVID-19 epidemic, using France as a test case and mortality data because of its extensive sampling compared to case incidence data.

## 2 Methods

### 2.1 The Discrete Stochastic (DS) model

Our model simulates the number of newly infected individuals per day (i.e. the daily incidence) as an iterative sequence following a Poisson distribution. We assume that the average number of secondary cases is equal to *R*_0_ and that the host population is homogeneously mixed (i.e. no spatial structure). These assumptions are relevant if a small fraction of the population is infected (Trapman et al., 2016).

More specifically, each individual is assumed to cause a random number of secondary infections throughout his/her infection, depending on his/her infectiousness (*β*). Here, infectiousness represents the relative infectious contact rate of an individual. It summarises both biological aspects (efficiency of transmission per contact, susceptibility of the recipients), and the contact rate of the individual, during the whole infectious period. Secondary infections occur randomly several days after contracting the disease. The probability of infecting someone some days after getting the disease is captured by the generation time, which we approximate using the serial interval (Nishiura, Linton, et al., 2020b).

Let *ω*_*a*_ be a random variable describing the probability of infecting someone *a* days after contracting the disease. An individual infected since *a* days infects new individuals at a rate *R*_0_ × *β* × *ω*_*a*_ during that day. Therefore, the number of secondary infections occurring *a* days after being infected, which is considered as a count of independent events, follows a Poisson distribution parameterized by *R*_0_ × *β* × *ω*_*a*_. From the additive property of the Poisson distribution, we find that the mean number of secondary cases during the entire infectious period is equal to the individual infectiousness. We then repeat this process for all individuals to determine the disease global progression.

Let *Y*_*t*_ be the random variable describing the incidence, i.e. the number of new infections, on day *t, t* being the number of days since initialisation of the process. The sequence of *Y*_*t*+1_ is defined using the Poisson additive property:

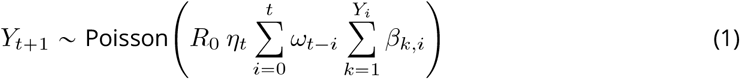

where *η*_*t*_ is the average normalized contact rate in the population at day *t, β*_*k,i*_ is the infectiousness of individual *k*, infected at day *i*, and *ω*_*t−i*_ is the probability of an individual infected at time *i* to infect someone at time *t* (*t − i* is the age of the infection).

We consider two scenarios (a) without and (b) with individual heterogeneity. If we denote by *ℬ* the distribution of random variables *β*_*x,y*_, accounting for the infectiousness of an individual *x* infected at day *y*, then, in each scenario we assume that:

a. *ℬ* is a Gamma distribution with shape parameter *k* = 0.16 and mean *R*_0_, implying that individuals are heterogeneous in infectiousness and/or contact rate, which can lead to ‘super-spreading’ events. We use the shape parameter (*k*) value estimated for a SARS outbreak in 2003 (Lloyd-Smith et al., 2005), which is consistent with estimates for SARS-CoV-2 epidemics (Althouse et al., 2020; Endo et al., 2020; Liu et al., 2020; Sun et al., 2020).
b. *ℬ* is a Dirac distribution, noted *δ*(1), implying that there is no heterogeneity and individuals have the same infectiousness and infection duration distribution. This is equivalent to *k →* + ∞ in the previous scenario. The sequence (*Y*_*t*_)_*t∈N*_ then simplifies into:

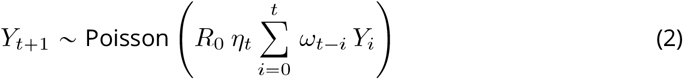

To model the control intensity over the epidemic at time *t* such as, for instance, a national lockdown, we vary the contact rate parameter *η*_*t*_. We assume that *η*_*t*_ is piecewise constant and that its discontinuities capture changes in public health policies (see Figure S1).

Overall, we define the temporal reproduction number (*R*_*t*_) at time *t* such that

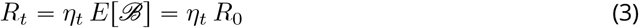

### 2.2 Beginning of the epidemic wave

To infer the starting date of the epidemic wave, we run our discrete stochastic (DS) algorithm starting from one infected individual until the infection dynamic becomes deterministic, *i*.*e*. the law of large numbers applies. We set the mortality incidence threshold to 100 daily cases, which was reached on 23 March 2020 in France. Neglecting the delay from infection to death, this would correspond to a daily incidence of more than 11,000 new cases according to the infection fatality ratio; a value much higher than the outbreak threshold above which a stochastic fade-out is unlikely (Hartfield and Alizon, 2013). We use independent estimates for the other parameters and perform a sensitivity analysis, shown in the Appendix.

To simulate death events in the DS model, we use the infection fatality ratio *p* (Verity et al., 2020), *i*.*e* the proportion of those infected who will go on to die from that infection. If we write *X*_*t*_ the number of individuals infected at time *t* who will die:

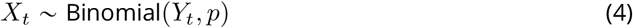

For each of the *X*_*t*_ individuals, the day of death is set by drawing a time from infection to death following *θ, i*.*e*. a Gamma distribution. *θ* was previously estimated on French hospital data (Sofonea et al., 2020) (Table S1), and its estimate compare very well with other independent estimates made from contact tracing data (Linton et al., 2020).

We repeat the algorithm 10,000 times to obtain a stable distribution of starting dates and discard epidemics that die out before reaching the threshold incidence. To allow for comparison with empirical data, we compute a sliding average of this time series over a 7-days window.

Finally, we assume that the consequences of the lockdown, which was initiated in France on March 17, did not affect the death incidence time series until the very end of March because of the delay between infection and death, which we estimate in France to be more than 11 days for 95% of the cases (Sofonea et al., 2020).

### 2.3 End of the epidemic wave

Here, we estimate how many additional days of lockdown would have been necessary to reach epidemic extinction for various lockdown intensity. Using the case of France as an example, the estimated lockdown contact rate is *η*_FR_ = 0.243, and we start our simulation on May 11, when the lockdown measures were partially lifted (*i*.*e* 55 days of lockdown). To avoid the accumulation of uncertainties, we initialise the model with incidence values obtained from a discrete-time non-Markovian model (Sofonea et al., 2020) for the past 15 days before the start of the simulation, in France. This threshold arises directly from the choice of the serial interval distribution: 99.9% of the transmissions occur within less than 15 days, using the generation time (Table S1).

We then use a Monte-Carlo procedure to estimate key features of the time series (*Y*_*t*_)_*t*_, such as the mean extinction time or the cumulative extinction probability. This is done by running 10,000 independent and identically distributed simulations of our process for each set of parameters.

We analyse the 10,000 resulting trajectories as follows. First, we estimate the distribution of *τ*, which is the random variable corresponding to the minimal lockdown duration (in days) such that the incidence is always null afterward for various scenarios. To mimic what happened during the first lockdown we set the contact rate to *η*_FR_ for the first 55 days. We then set the contact rate to a fixed value (greater or equal than *η*_FR_) until extinction is reached. As long as the effective reproductive number is lower than 1, the time to extinction is finite. Mathematically,

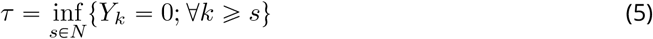

Second, we study the effect of finite lockdown extensions on the probability of extinction to understand the risk of epidemic rebound upon lockdown lifting. For simplicity, we assume that control measures are completely lifted once the lockdown is over. The probability of having no new cases at time *t* (*p*_0_(*t*)) is estimated using the following formula

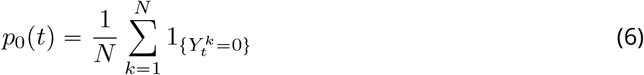

where *N* is the number of simulations performed and 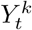 the number of newly infected individuals in the *k*^th^ simulation at time *t*.

Third, we study the effect of initiating the first lockdown one month or two weeks earlier (in France, on February 17 or March 03 respectively) on the distribution of the time to extinction (*τ*). For comparison purposes, we assume that in any case the first 55 days of lockdown have the same contact rate (*η*_*t≤*55_ = *η*_FR_) and then extend the lockdown indefinitely with variable intensities to estimate the time to extinction (*τ*) as described previously (equation 5).

### 2.4 Alternative models

To further study the effects of stochasticity, non-Markovian dynamics, and superspreading, we implemented two deterministic models. The first is Markovian, i.e. memoryless, and is based on a simpler model derived from a classical SEIR model. The second has a discrete-time structure, which allows capturing non-Markovian dynamics (Sofonea et al., 2020).

#### The SEAIRHD model

In this classical compartment model, hosts can belong to seven states: susceptible to infection (*S*), exposed (i.e. infected but not infectious, *E*), asymptomatic and infectious (*A*), infectious and symptomatic (*I*), removed (i.e. recovered or isolated, *R*), hospitalised who will die (*H*), or dead (*D*) (Fig. S2). The model is described by a set of ODE detailed in the Appendix (equation system **??**). Since the model is deterministic, we can seed the simulations with a single exposed individual on day *t*_0_.

This model is solved numerically using the Numpy package in Python 3.8.3 to obtain a deterministic trajectory. Parameters were chosen with maximal likelihood given the observed daily mortality data, assuming that the daily mortality incidence is Poisson distributed, and independence between daily incidences (For more details, see the supplementary material). We also simulate a stochastic version of this model 1,000 times using a Gillespie algorithm with the package TiPS (Danesh, Saulnier, et al., 2020) in R v.3.6.3 R Core Team, 2020.

#### COVIDSIM: A non-Markovian deterministic model

Finally, we use an existing discrete-time model that has a similar structure to the continuous model mentioned above with an additional age-structure (Sofonea et al., 2020). For comparison purposes, the generation time is set to be the same as in our DS model (Nishiura, Linton, et al., 2020a), and so the (non-exponential delay) from infection to death. However, two major differences are that this third model is not stochastic and does not allow for superspreading events. We restricted the parameter inference to the daily hospital mortality data described previously, with the main free parameter being the date of origin. We invite the reader to refer to Sofonea et al., 2020 for the scripts and further details on this approach.

### 2.5 Model calibration

To allow for model comparison and improve estimates, we fix some key parameters based on existing values, focusing on the French COVID-19 epidemic. Table S1 lists all the parameters used along with key references.

We compute the likelihood of the deterministic SEAIRHD model assuming a Poisson distribution of the daily mortality incidence data. Parameter inference with maximum likelihood is performed using the Nelder-Mead algorithm implemented by Scipy.minimize function in Python.

The parameters used for the non-Markovian deterministic model correspond to the maximum likelihood set of parameters used in Sofonea et al., 2020.

### 2.6 Code and simulation results availability

The different scripts and simulation results are available on Gitlab: https://gitlab.in2p3.fr/ete/origin-end-covid-19-epidemics

## 3 Results

### 3.1 Origin of the epidemic wave

When neglecting host heterogeneity, using our DS framework, the median delay between the importation of the first case of the epidemic wave and the time mortality incidence reaches 100 deaths per day (March 23) is 67 days (equivalent to a first case on January 16 in France), with a 95% confidence interval (95% CI) between 62 and 79 days, *i*.*e*. between January 4 and 21 in France (Fig. 1). With this model, only 7% of the outbreaks die out before reaching the threshold.

**Figure 1.**
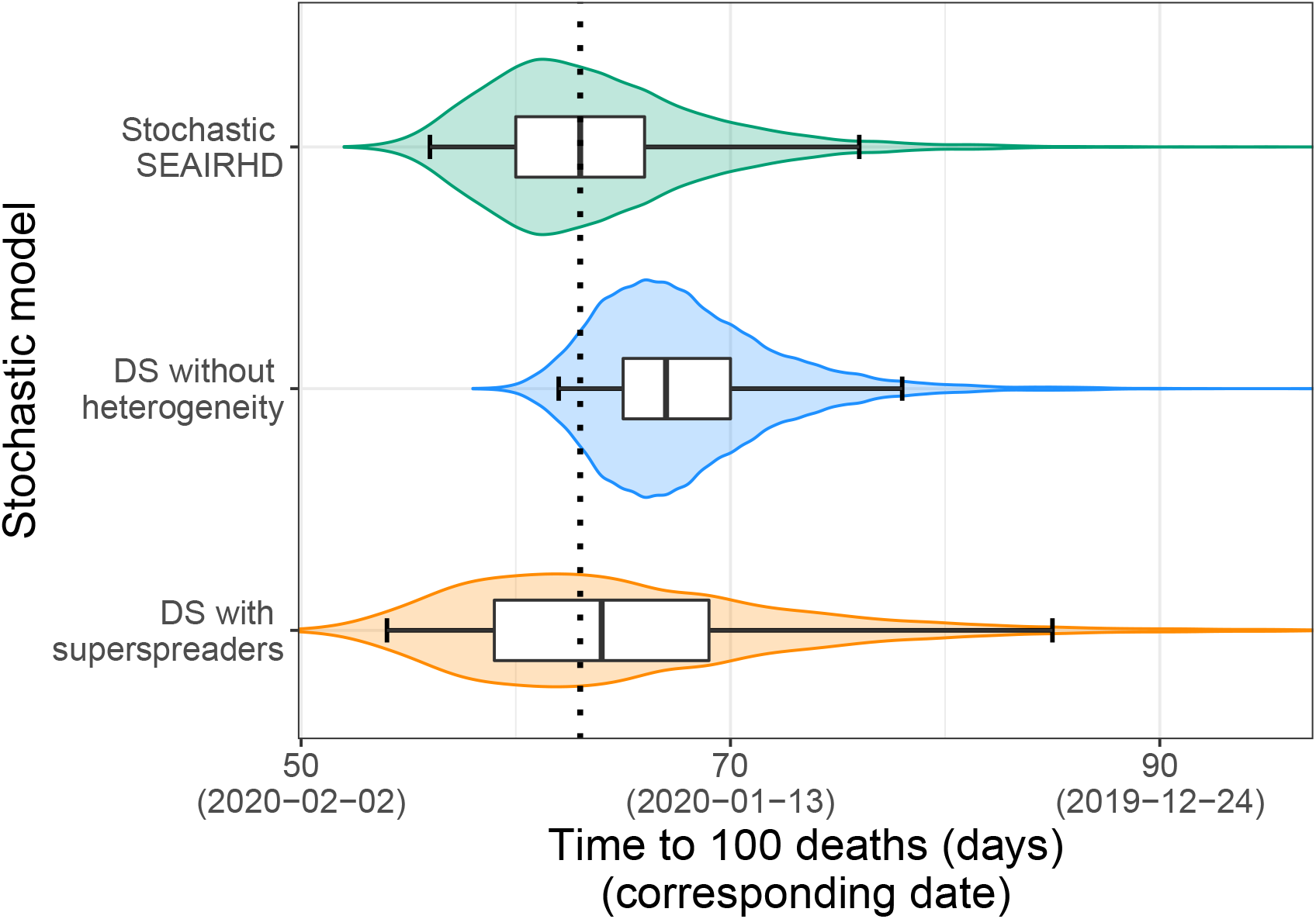
Estimated distribution of the number of days until daily mortality incidence reaches 100 cases. The boxplots and the whiskers indicate the 2.5%, 25%, 50%, 75%, and 97.5% quantiles out of the 10,000 simulations. The red dashed line shows the estimates using the deterministic models.

Superspreading events, *i.e*. when the individual infectiousness *ℬ* follows a Gamma distribution, seem to have limited effects on these results: the median delay drops slightly to 64 days (January 19 in France), although with a larger 95% CI, between 54 and 85 days. Moreover, as expected (Lloyd-Smith et al., 2005), we observe a soar in the frequency of epidemic outbreaks dying out before reaching the threshold, which represent 75% of our simulations.

When assuming deterministic and Markovian dynamics with our SEAIRHD model, the importation date of the first case of the epidemic wave that best fits the results is similar, with a delay of 63 days until daily mortality incidence reaches 100 cases. A stochastic implementation of the same model yields the same median delay of 63 days [95% CI: 56 - 76 days], which is comparable to the DS model. However, consistently with earlier studies (Grant, 2020; Sofonea et al., 2020), the ability of this memoryless model to capture the data is limited (Fig. S3 in the Appendix). Finally, the maximum likelihood parameter estimates from a deterministic but non-Markovian model, COVIDSIM (Sofonea et al., 2020), restricted to the mortality data, indicates a similar delay of 63 days (January 20), [95% CI: 63 - 64 days].

We perform a sensitivity analysis of our results focusing on two parameters. First, we show that the median delay for daily mortality to reach 100 cases is increased by 5 days when the generation time standard deviation is decreased by one third (Fig. S5). Therefore, the estimates remain within the confidence interval obtained for the starting date of the epidemic. Second, increasing the number of initially imported cases from 1 to 5 decreases the delay by 7 days, with a median of 60 days [95% CI: 57-64 days] without heterogeneity. However, when assuming a more realistic scenario where all those cases are not imported on the same day, this impact of the delay was more limited (Fig. S6). For example, if the 5 cases are imported during the first five days of the outbreak, the decrease is only of 5 days, with a median delay of 62 days [95% CI: 59 - 66 days].

Overall, non-Markovian dynamics or stochasticity do not tend to strongly impact the estimate of the delay for an epidemic to reach a daily mortality incidence of 100 cases. Introducing super-spreading events, however, slightly decreases the delay estimated and greatly increases its variance. As expected, the initial number of imported cases can have an impact on the estimates.

### 3.2 End of the epidemic wave with lockdown

#### Time to eradication

We estimate the distribution of the minimal lockdown duration to eradicate the epidemic (*τ*) by first neglecting superspreading events and starting from the end of the first-wave lockdown in France on May 11 (orange violins in Figure 2). When maintaining the constraints on social interactions to their full intensity (*η*_*t* > 55_ = 0.24), a total of at least 8 months of lockdown, including the 55 days between Mar 17 and May 11, are required to reach a 97.5% extinction probability.

**Figure 2.**
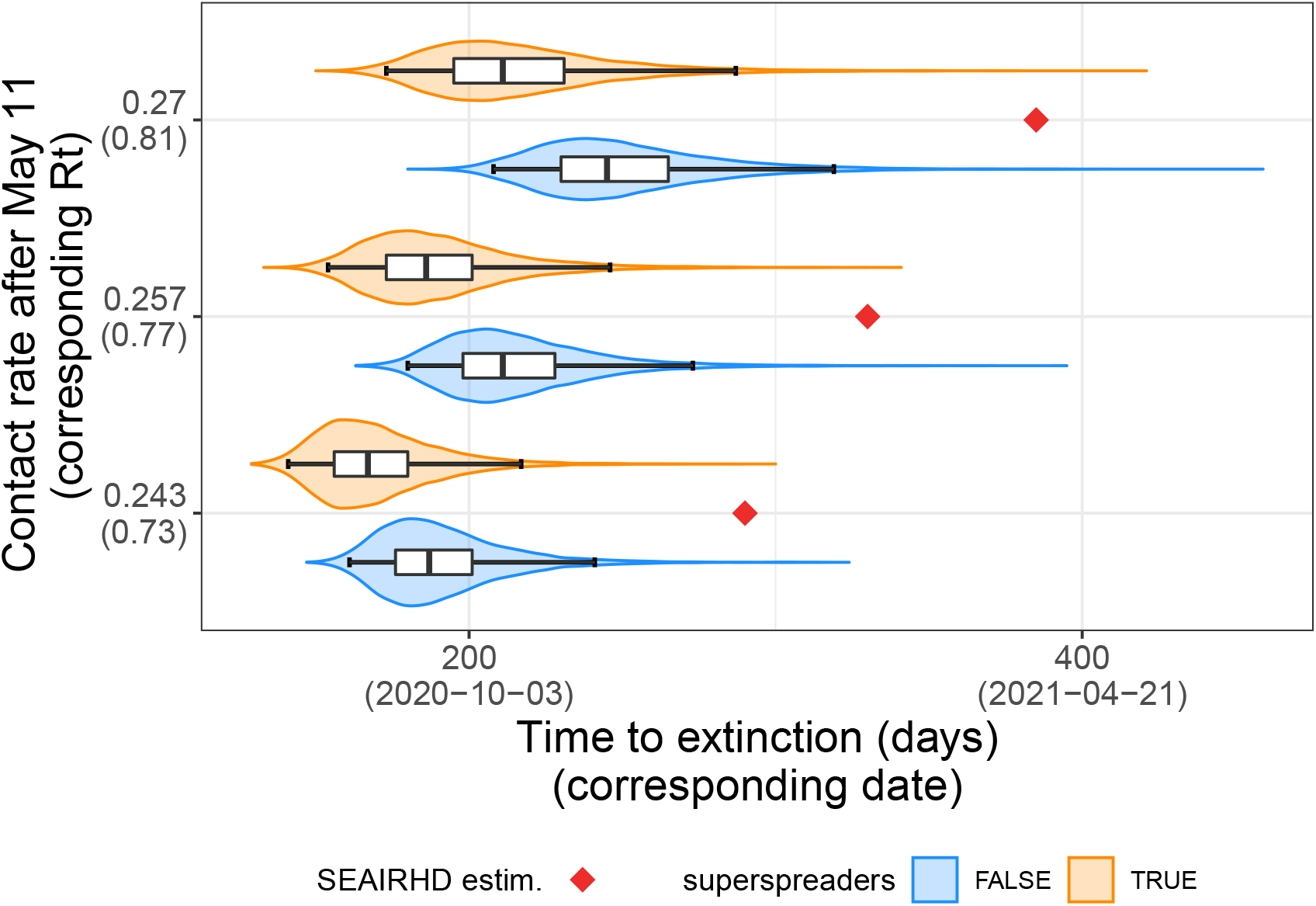
Effect of lockdown intensity, stochasticity, and superspreading events on the time to extinction (*τ*). The distributions of the time to extinction (in days since the start of the lockdown on March 17) for several lockdown intensities (*η*_*t*_) after the first 55 days (i.e. after 11 May 2020) are plotted on the Y-axis using violin plots and boxplots. Results without transmission heterogeneity (*ℬ*= *δ* (*R*_0_)) are in orange. In blue, we assume a Gamma distribution for *ℬ*. Red diamonds show results from the deterministic Markovian model. The box extends from the lower to upper quartiles of the data. The whiskers expand from the 2.5% to the 97.5% quantiles.

When accounting for individuals heterogeneity, we find that, everything else being equal, the quantiles of the time to eradication (*τ*) are always lower than the homogeneous cases. However, 7.23 months of lockdown at full intensity (*η*_*t*>55_ = 0.24) are still required to guarantee extinction in 97.5% of the cases (blue violins in Figure 2). Accounting for individual heterogeneity also reduces the variance of *τ*. This is expected because in this case, the majority of the infected people do not transmit, which increases the extinction probability (Lloyd-Smith et al., 2005).

The mean values of the time to eradication (*τ*) increases with the decrease in the intensity of the lockdown constraints post 55 first days of lockdown. As the contact rate of the population tends towards 1{*R*_0_ the mean values of *τ* diverge towards infinity. The dynamical process is said to be critical (resp. super-critical) if *η*_*t*_ *=* 1/*R*_0_ (resp. *η*_*t*_ ⩾ 1/*R*_0_). This result holds when assuming transmission heterogeneity.

We also compute the time to extinction with the deterministic SEAIRHD model after tuning the model using the parameters that best fitted the mortality incidence (Fig. 2). The time to extinction corresponds here to the minimum time where the incidence reaches zero.

#### Rebound risk

In our stochastic model, the incidence at time *t* (denoted (*Y*_*t*_)_*t ∈N*_) can alternate between zero and non-zero values. To evaluate the risk of epidemic rebound, we implement a finite lockdown extension after which all constraints are released (*η*_*t*_ = 1). This allows us to calculate *p*_0_(*t*), the probability to have 0 new cases after time *t*. In Figure S7, we see a sharp decrease in *p*_0_(*t*) a few days after lockdown release.

The rebound risk is directly linked to the transmission heterogeneity. Assuming a higher individual transmission heterogeneity (i.e. lower *k*) drastically reduces the risk of rebound, as it also implies that most infectees do not transmit the disease.

#### Eradication and lockdown initiation date

We now turn to the consequence of implementing a lockdown a month or two weeks earlier. In France, this corresponds to Feb 17 and Mar 03 (at that time, a total of respectively 1 and 3 deaths were reported).

The results are shown in Figure S8 for the case without host heterogeneity and Fig. 3 with superspreading events. Initiating the lockdown one month earlier, i.e. for France approximately 33 days after the onset of the epidemic wave, decreases the 97.5% quantile of the time to extinction by 91 days with transmission heterogeneity (97 days without heterogeneity) in the most restrictive scenario. If the onset of the lockdown is brought forward by two weeks (March 3^rd^), *i.e*. in France approximately 48 days after the onset of the epidemic, 97.5% of the extinction events occur before the 178^th^ day of lockdown with transmission heterogeneity (199^th^ day without heterogeneity). Hence a reduction of 39 (resp. 42) days of lockdown could be expected compared to the actual start (Mar 17).

**Figure 3.**
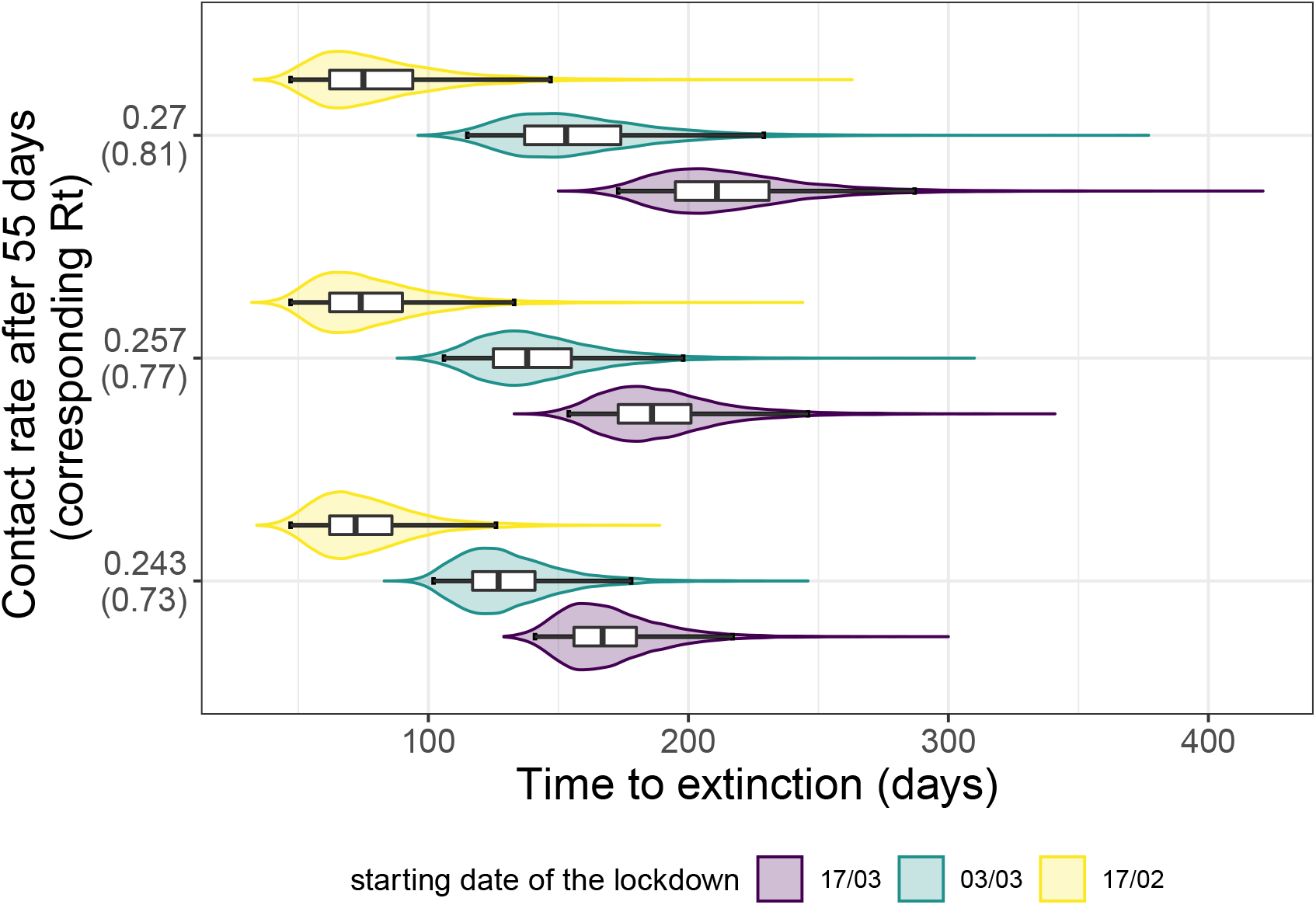
Effect of the lockdown intensity, stochasticity, and initiation date on the time to extinction (*τ*) under individual spreading heterogeneity assumption. The distributions of the time to extinction (in days since the start of the lockdown) for several contact rate restrictions post 55 first days are plotted on the Y-axis using violin plots and boxplots. In this graph, we assume individual spreading heterogeneity. The colors indicate the different initiation date of the lockdown: in purple it starts on Feb 17, green Mar 03, and yellow on Mar 17 (official start). The box extends from the lower to upper quartiles of the data. The whiskers expand from the 2.5% to the 97.5% quantiles.

These numbers increase with the easing of the constraints following the first 55 days of strict lockdown (*η*_*t*_ = 0.24). When assuming a lighter control in the following days (e.g. *η*_*t ≥*55_ = 0.29), one can notice that the increase in the quantiles of *τ* when starting the lockdown on Feb 17 is much lower than the two other cases.

#### Time to a threshold of 20 new cases per day

Finally, we study the distribution of the delay to reach 20 new cases per day, below which it is expected that a general lockdown is not required to control the epidemic. We evaluate the effect of lockdown intensity, initiation date and individual spreading heterogeneity on this delay.

The estimated distributions of the time to 20 new daily cases when accounting for superspreading events is displayed in Figure 4 (see Figure S9 for the estimations without superspreaders). Our model suggests that initiating control measures one month earlier (mid-February) would have reduced the 97.5% quantile of the time to 20 new cases by 95 days under the strictest restrictions. In the mid-February scenario, we notice the time to 20 new cases occurs during the 55 first days of lockdown. Starting the lockdown early March does reduce the 97.5% quantile of the time to the threshold by 40 days. However, the first 55 days of lockdown are not sufficient to reach the limit of 20 new cases per day.

**Figure 4.**
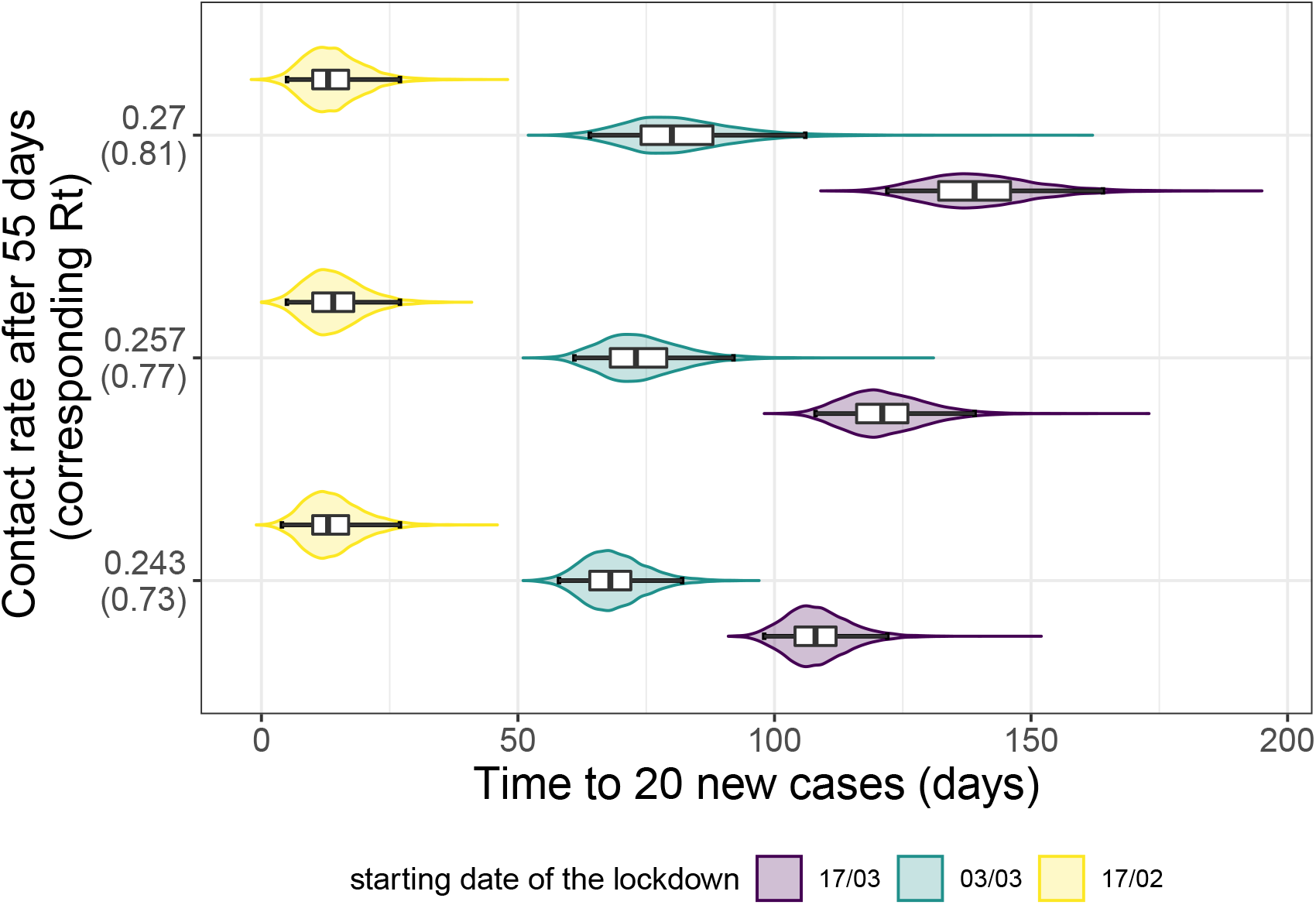
Effect of the lockdown intensity, stochasticity, and initiation date on the time to 20 new cases under individual spreading heterogeneity assumption. The distributions of the time to 20 new cases (in days since the start of the lockdown) for several contact rate restrictions post 55 first days are plotted on the Y-axis using violin plots and boxplots. In this graph we assume individual spreading heterogeneity. The colors indicate the different initiation date of the lockdown: in purple it starts on Feb 17, green Mar 03 and yellow on Mar 17 (official start). The box extends from the lower to upper quartiles of the data. The whiskers expand from the 2.5% to the 97.5% quantiles.

## 4 Discussion

In the early and final stages of an epidemic, stochastic forces may strongly affect transmission dynamics because infection prevalence is low. Using stochastic mathematical modelling, and assuming *R*_0_ = 3, we estimate the time for a COVID-19 epidemic to reach an incidence of 100 deaths per day to be approximately 67 days, with a 95% probability between 62 and 79 days. In the case of France, where such incidence values were reached on Mar 23, this translates into an origin of the epidemic around January 16, with 95% probability between January 4 and 21. This is consistent with estimates obtained using virus genome data, although these should be interpreted with caution due to the uncertainties regarding the molecular clock estimates for the virus and the incomplete sampling in France (Danesh, Elie, et al., 2020).

Accounting for superspreading events yields a later median date of origin (January 19 for France). This is expected because, in outbreaks that do not die out, superspreading events accelerate the initial dynamics (Lloyd-Smith et al., 2005). However, this difference is not significant.

The 95% CI for the epidemic starting date generated by our different models overlap. This could originate from our use of mortality data. Since death occurs after a mean delay of 23 days after infection (Sofonea et al., 2020), by the time mortality incidence is detectable, transmission dynamics are largely deterministic. This also explains why introducing superspreading events mostly affects the variance of the estimate. Unfortunately, hospital admission data is not available for France until 18 March 2020, and screening data was initially performed with a very low sampling rate in the country (only severe cases were tested).

Care must be taken when comparing the estimates from our discrete stochastic model to that of earlier models. For instance, the non-Markovian deterministic COVIDSIM model (Sofonea et al., 2020), which estimates the date of onset to be slightly later (January 20), includes host age structure. Regarding the more classical deterministic and Markovian SEAIRHD model, its ability to fit the data is limited (Fig. S3), except when only considering the exponential phase before the lockdown. This poor inference of underlying epidemiological dynamics is likely due to the absence of memory in the underlying processes, as stressed by earlier studies (Grant, 2020; Sofonea et al., 2020). When incorporating memory on the hospitalization-to-death delay, we obtain a much better fit, and the time to the daily mortality of 100 cases is then comparable to that of the model without superspreading events.

We also estimate the median number of days of full intensity lockdown required to achieve extinction with a 95% confidence. In the French setting (i.e. introduction of the lockdown after 67 days of the epidemic), we find with our stochastic model that 187 (95% CI: [161, 241]) days of lockdown would be required to reach extinction in a homogeneous transmission scenario in 50% of the cases. Accounting for superspreading events decreases the median estimate value by 20 days. Initiating the lockdown one month earlier strongly affects these estimates: a 30 days anticipated start reduces the mean number of days spent in full intensity lockdown by 95 days, i.e. a 51% reduction.

50% of the simulations reach the threshold of 20 new cases after 108 (95% CI: [98, 122]) days of lockdown at full intensity initiated mid-March. When initiating the constraint in mid-February, this threshold is reached in 13 (95% CI: [4, 27]) days. Since, in the latter scenario, the epidemic spread is more limited, the first 55 days of lockdown are decisive in the slowing down of the epidemic. This confirms that early interventions have a disproportionate impact on the epidemic dynamic.

Finally, we investigated the risk of an epidemic rebound upon lockdown lifting. In this scenario, super-spreading has a striking impact in limiting this risk, which is consistent with earlier work on outbreak emergence (Lloyd-Smith et al., 2005).

There are several limitations to this work. First, the generation time *ω* and the time from infection to death *θ*, remain largely unknown in France, as well as in many countries. Most serial interval estimates rely on contact tracing data from Asia (Liu et al., 2020; Nishiura, Linton, et al., 2020b), which could differ from the distribution in France, due to differences in contact structure, or non-pharmaceutical interventions. Although the generation time distribution is expected to affect epidemic dynamics, we show in Figure S4 that the variance of this interval has little impact on our results.

Another important limitation about the estimation of the date of origin of the epidemic comes from the assumption that only a single infected person caused the epidemic. Most epidemics outside China were seeded by multiple importation events. The problem is that there is an identifiability issue because it is impossible to estimate both the number of initial infected cases and the time to a threshold of 100 deaths with incidence data only. However, some estimates made in the UK from phylogenetic data as well as the combination of prevalence and travel data show that the estimated number of importation events is less than 5 per day before the end of February, when the virus was beginning to circulate at higher levels throughout Europe (Plessis et al., 2021). Assuming a similar importation pattern in France, we show that the dynamic is only sensitive to the importation events within the first days after the beginning of the epidemic wave. While these events may have helped the epidemic to escape the stochastic phase faster, they are unlikely to strongly affect the estimated date of the beginning of the wave (Figure S6). In a quite extreme scenario of 5 importations per day during 30 days, we estimate the median day of the epidemic beginning to be 16 days later (*i.e*. Feb 2 for France).

Another limitation comes from the lack of data regarding individual heterogeneity in COVID-19 epidemics. Such heterogeneity was supported by early limited data (Endo et al., 2020; Liu et al., 2020) but recent additional evidence from Chinese transmission chains further supports this result (Sun et al., 2020), although with a higher *k* parameter value than the one used here (0.30 versus 0.16 here), meaning a less heterogeneous transmission. Therefore, our assessment of superspreading events impact seems conservative.

These results have several implications. First, they can help reconcile the fact that cases may be detected long before the emergence of the transmission chains related to an epidemic wave. This is particularly important for an audience not familiar with stochasticity. Second, the estimate of the time required to ensure that the epidemic is gone can help inform public health decisions. In the case of the French epidemic, for instance, enforcing a strict lockdown from March 17 until epidemic extinction was practically unfeasible. However, this may not be the case if measures are taken early enough in the epidemic. Furthermore, our work also illustrates the risk of epidemic rebound as a function of the duration of the lockdown. Overall, this work calls for further studies, especially to assess the importance of super-spreading events in the global spread of SARS-CoV-2.

## Data Availability

Codes and results are available on gitlab.

https://gitlab.in2p3.fr/ete/origin-end-covid-19-epidemics

## Acknowledgements

Thomas Beneteau is supported by a doctoral grant from the Ligue Contre le Cancer. We thank the CNRS, the IRD, and the IRD itrop HPC (South Green Platform) at IRD montpellier for providing HPC resources that have contributed to the research results reported within this paper. We also thank the ETE modelling team for discussion. The authors of this preprint declare that they have no financial conflict of interest with the content of this article. This article was submitted on medRxiv (Bénéteau et al., 2021). Version 3 of this preprint has been peer-reviewed and recommended by *Peer Community In Mathematical and Computational Biology* (https://doi.org/10.24072/pci.mcb.100004).

## Appendix

### Models parameters

**Table S1.** Models parameters and constraints.

| Notation | Signification | Model | Constraint/value | Reference |
| --- | --- | --- | --- | --- |
| $R_0$ | Basic reproduction number | all | 3.02 | Sofonea et al., 2020 |
| $p$ | Infection fatality ratio | all | 0.874 % | Sofonea et al., 2020 |
| $\eta$ | Contact rate in the population | all | 0.243 | inferred |
| $(\omega_i)_{i \in N}$ | Serial interval distribution | stochastic | discretised *Weibull* (mean: 4.8 d., sd: 2.3 d.) | Nishiura, Linton, et al., 2020b |
| $\theta$ | Infection to death delay | stochastic | *LogNormal*(mean: 23.3 d., sd: 9.7 d.) | Sofonea et al., 2020 |
| $k$ | Shape parameter (Gamma distribution of the infectivity) | stochastic | 0.16 | (Lloyd-Smith et al., 2005) |
| $t_0$ | Time of epidemic wave initiation | SEAIRHD | 2020/01/22 | inferred |
| $\beta$ | *Per capita* infectious rate | SEAIRHD | $\frac{R_0 \gamma \sigma}{S_0 (\gamma + \sigma)}$ | calculated |
| $\epsilon$ | Rate of end of latency | SEAIRHD | 0.415 | inferred |
| $\sigma$ | Rate of symptoms onset | SEAIRHD | $\frac{1}{5.6 - 1/\epsilon}$ | Linton et al., 2020 |
| $\gamma$ | End of infectivity rate | SEAIRHD | 0.653 | inferred |
| $\alpha$ | Death rate | SEAIRHD | $\frac{1}{17.7 - 1/\gamma}$ | Sofonea et al., 2020 |
| $\mathcal{N}$ | total population | SEAIRHD | 64171900 | INSEE |

#### Contact rate variation during and post lock-down period

**Figure S1.**
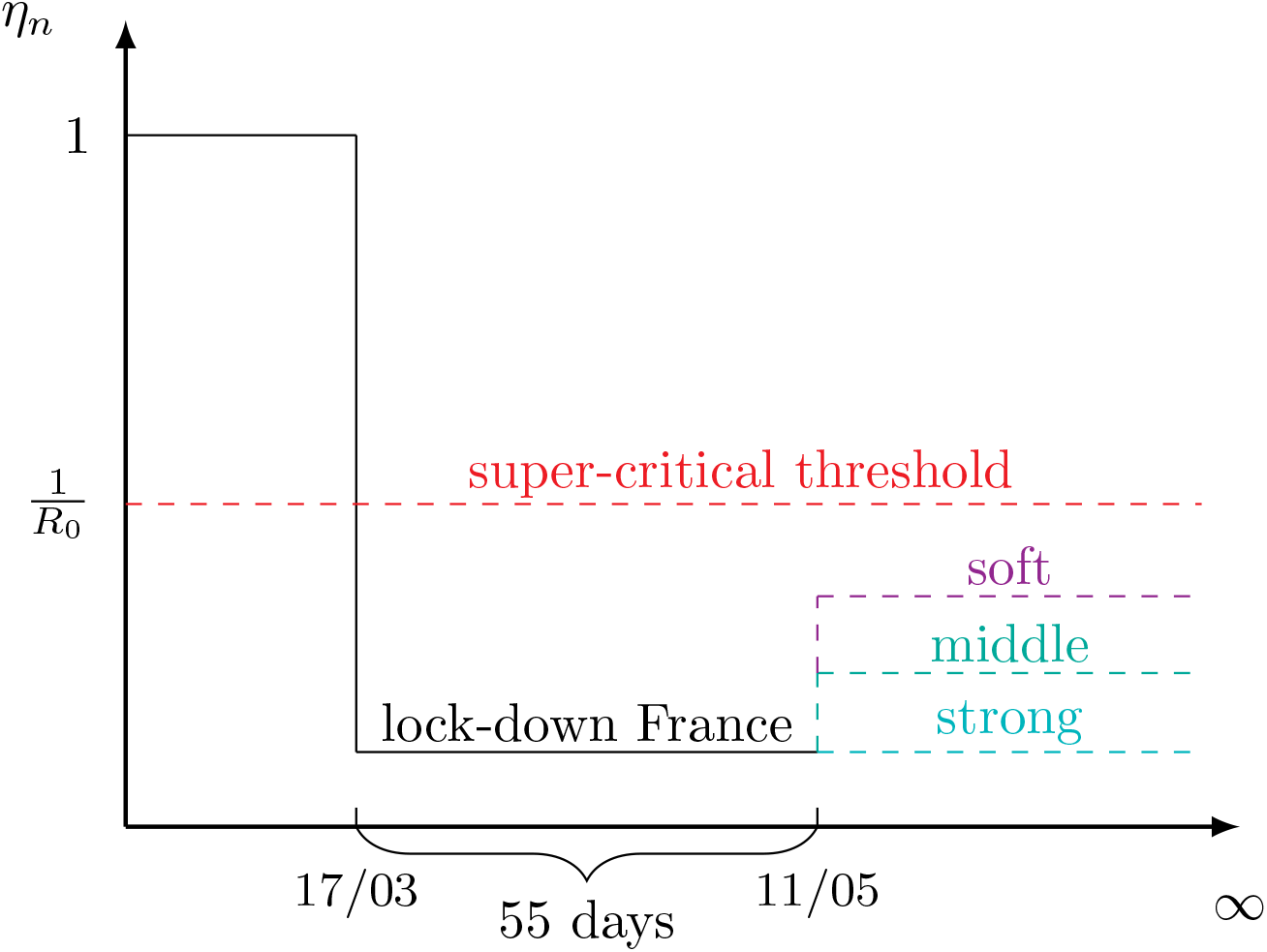
Variation of the contact rate (*η*_*t*_) for the estimation of the time to extinction (*τ*). On Mar 17 a lock-down was instated in France. We estimated its efficacy to be around 1-*η*_*t*_ = 0.76 (Sofonea et al., 2020). The lock-down officially ended on May 11, for a total of 55 days under full lock-down. We evaluated the time to reach extinction under various extension constraints by assuming an infinite lock-down prolongation with fixed intensity, potentially different from the value estimated during the Mar 17 and May 11. The intensity of the lock-down/extension is inversely proportional to the contact rate (*η*_*t*_).

#### Markovian SEAIRHD model

The model is captured by the following set of differential equations:

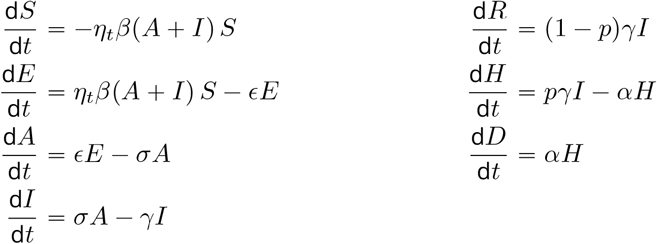

where *η*_*t*_ measures the public health intervention impacts on the disease spread at day *t, β* is the per capita transmission rate of asymptomatic and symptomatic hosts, is the rate of end of latency, *σ* is the rate of symptoms onset, *γ* is the recovery rate, *p* is the infection fatality ratio (IFR), and *α* is the rate at which hospitalised patients die. Our goal is to capture the key features of the infection life-history, especially the incubation period, the asymptomatic transmission, and the delay to hospitalized deaths, but not to fit the epidemic in details.

**Figure S2.**
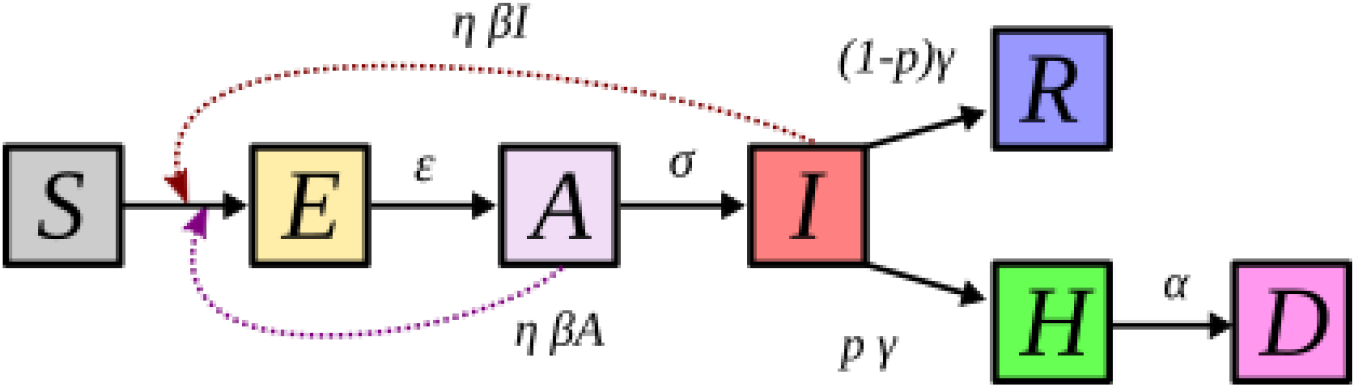
Flow diagram of the SEAIRHD model. This SEAIRHD model corresponds to the ODE model detailed in eq. S1a to S1g. Susceptible individuals (*S*) can be infected and become asymptomatically infected, but noninfectious (*E*). After a delay, they become asymptomatically infectious (*A*). After symptoms onset, they are symptomatic and infectious (*I*). A proportion will recover (*R*), with or without hospitalisation, but some, after a delay during which they are not infectious anymore (*H* compartment), will die (*D*). Parameter notations are shown in the main text.

##### SEAIRH4D model

An alternative model is the SEAIRH4D: in this case, memory is introduced on the delay from infection to death, *ie* it follows an Erlang with shape parameter 4 and the same mean. This is biologically more realistic than the SEAIRHD model where this delay is exponentially distributed.

##### Deterministic implementation

The set of ODE shown in the previous paragraph is solved using ‘odeint’ function from Numpy on Python 3.8.3. We then applied a moving average, with a window of 7 days, as done with the real data. We estimated the following parameters for the SEAIRHD and SEAIRH4D models using a maximum likelihood procedure: *t*_0_, *γ, σ* and *η*_*lockdown*_. The maximum-likelihood was found using the Nelder-Mead procedure implemented in the ‘minimize’ function from Scipy. We computed the likelihood assuming that the daily mortality incidence is Poisson distributed, and independence between daily incidences. We used the data of daily hospital mortality in France from January 1st to May 11 (end of national lockdown), on which a moving average of 7 days is applied to avoid “week-end effects”. The code is available on the gitlab repository (see main text Methods).

We compared those two models to the discrete time non-markovian model (Fig. S3).

**Figure S3.**
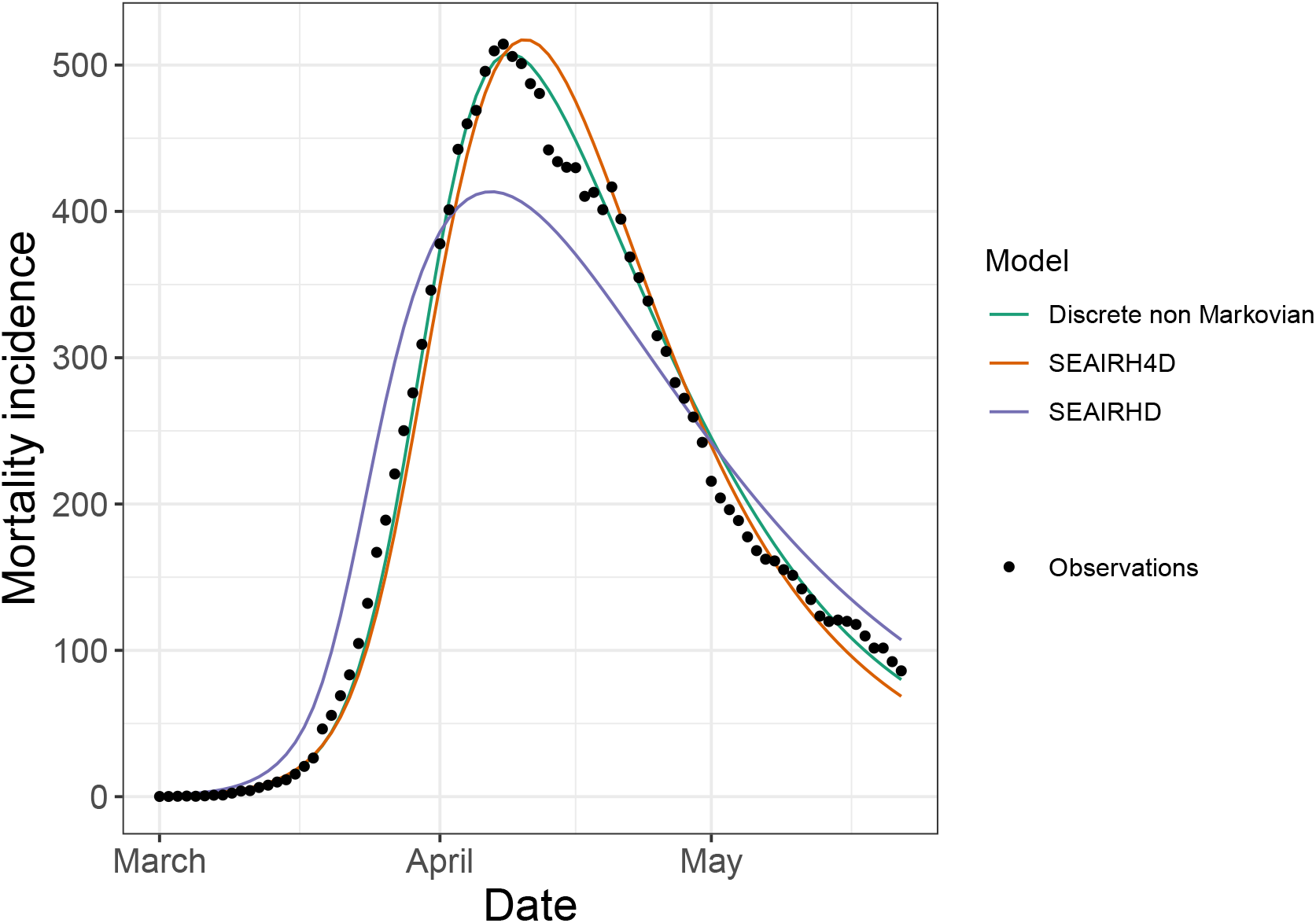
Best fits to the observed mortality incidence for several models. We estimated the best fits to the observed data by minimising the Akaike Information Criterion (AIC). The SEAIRHD shows mediocre fit to the data (AIC: 1324) but taking into account a more realistic delay from infection to death, SEAIRH4D model, improves the estimations drastically (AIC: 730). The discrete time model with memory effects provides a better fit (AIC: 461).

##### Stochastic implementation

Using the same parameters, we simulated 1,000 times a stochastic version of this model, using a Gillespie algorithm with the package *TiPS* (Danesh, Saulnier, et al., 2020) on R v.3.6.3 (R Core Team, 2020). We stored the time to 100 daily deaths, after applying a moving average with a window of 7 days on the simulations.

### DS model sensitivity analysis

#### Effect of the serial interval distribution

The time elapsed between infection events in an infector-infectee pair is called the generation time. It is a key epidemiological parameter in our model, though almost impossible to measure directly. However, the serial interval distribution - i.e. the time between symptom onsets in an infector-infectee pair-has theoretically the same expectation, but a higher variance.

**Figure S4.**
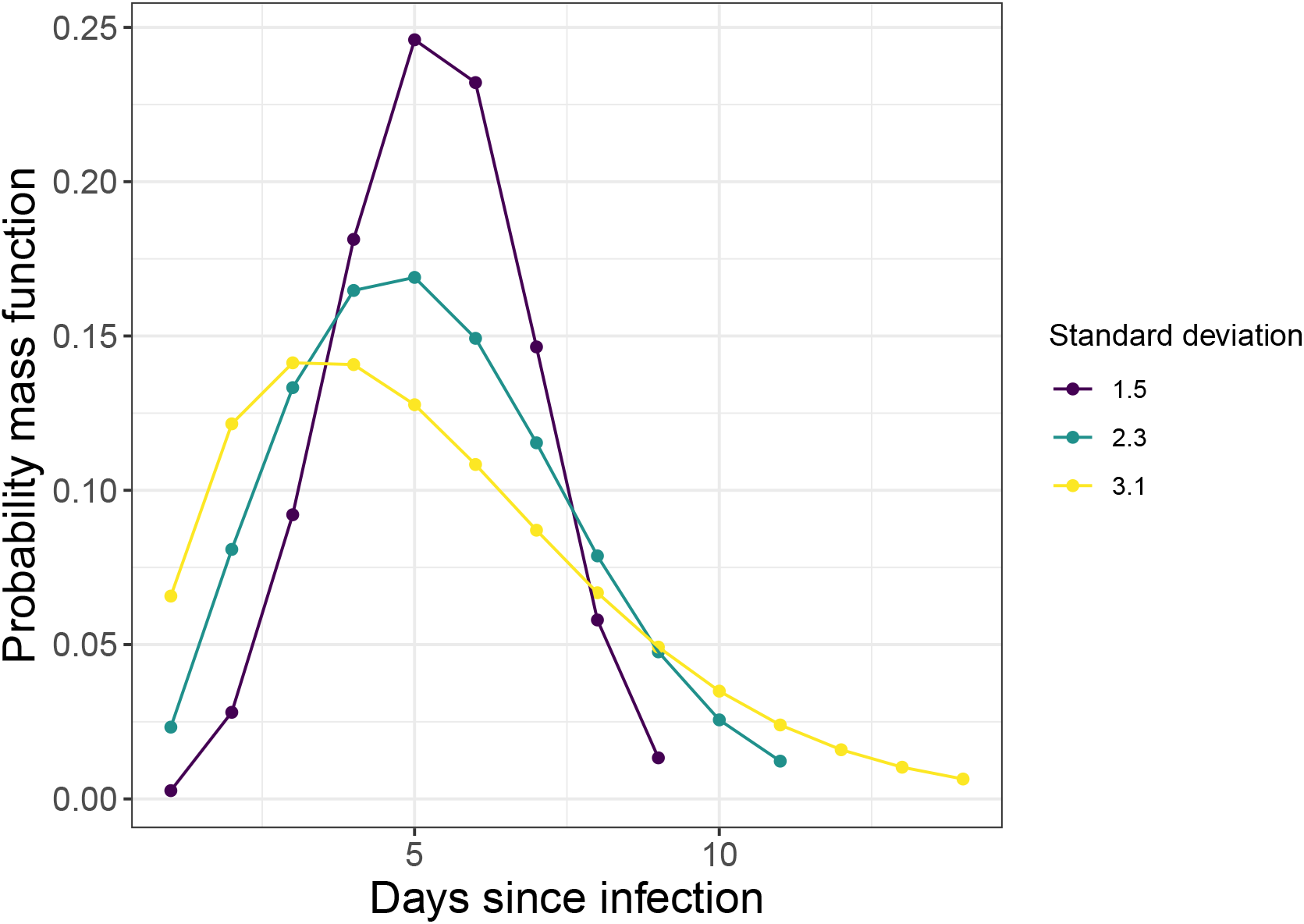
Probability mass function for Weibull distributions with mean 4.8 and various standard deviation. Serial interval distributions used hereafter to analyze the sensitivity to the serial interval standard deviation.

To our knowledge, the only available data to estimate this parameter come from Asia (Li et al., 2020; Nishiura, Linton, et al., 2020a). In our results, we use the distribution inferred by Nishiura, Linton, et al., 2020a, *i.e*. a Weibull distribution with mean 4.8 days and standard deviation 2.3 days. This estimate could change, e.g. through behavioral on contact structure between the countries where the data come from and France. Intuitively, the epidemic growth is very sensitive to the generation time expectation, hence the epidemic starting date would be shifted significantly. Here, we focus on the generation time variance to see to what extent the generation time distribution can affect the epidemiological dynamic (fig. S4).

We observe that the higher the standard deviation, the later the starting date is inferred (Fig. S5). The median value is shifted from January 11th to 19th when the standard deviation is doubled, under the homogeneous infectivity model. However, this variation remains within the interval containing 95 % of the variation, using the serial interval determined by Nishiura, Linton, et al., 2020a.

**Figure S5.**
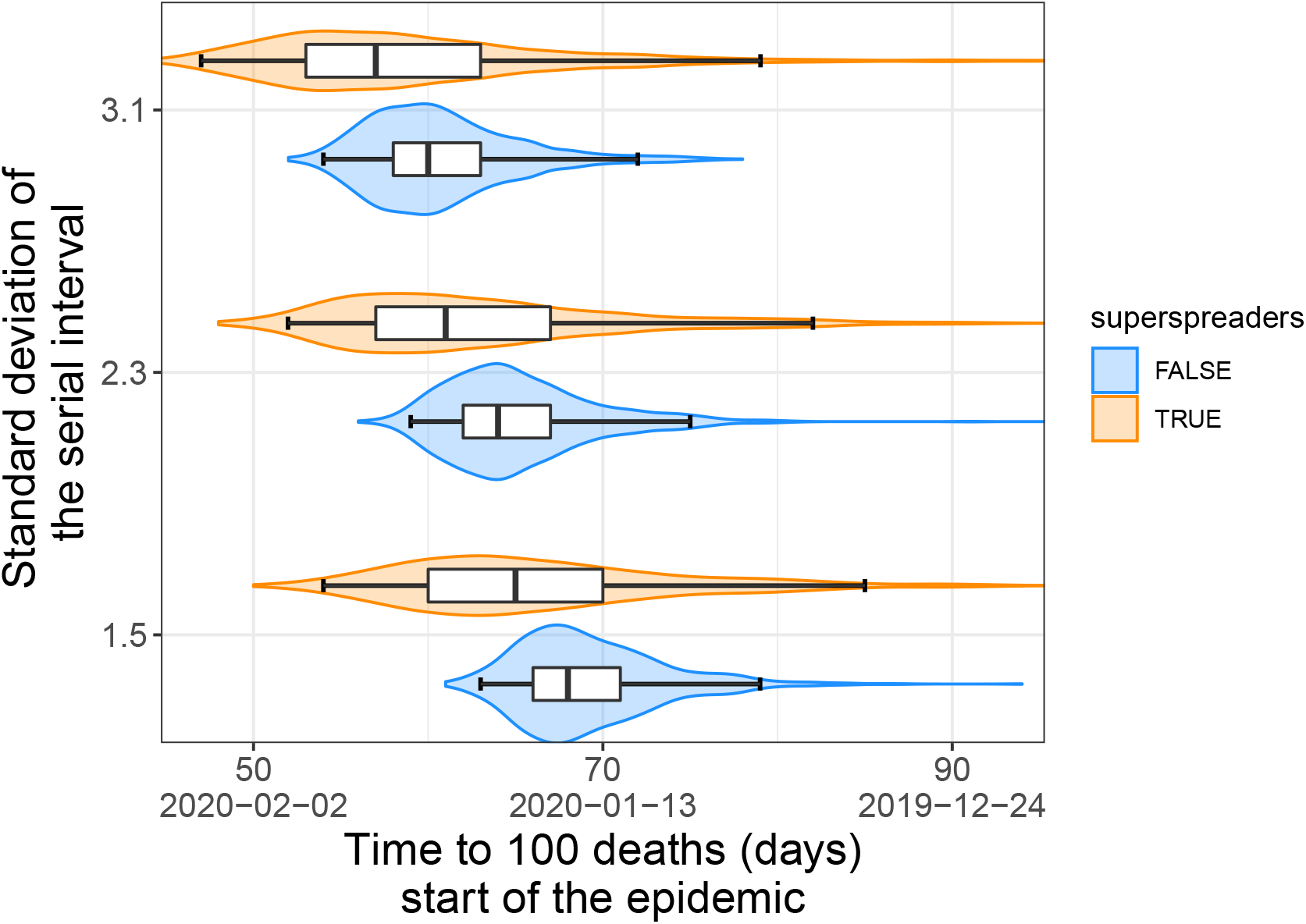
Time to 100 deaths distributions for various standard deviation of the serial interval distribution. We remind that all serial intervals share similar mean following (Nishiura, Linton, et al., 2020a) estimations. The boxplots show the following quantiles: 2.5%, 25%, 50 %, 75%, 97.5 %. An increase in the standard deviation of the serial interval distribution shorten the time to 100 deaths.

#### Effect of the initial number of imported cases

We made the assumption that only one imported case was responsible for the whole outbreak wave. To note, this is not incompatible with the occurrence of earlier cases, such as the one that occurred on December 27th in France, because in case of individual *R*_0_ heterogeneity, most of those early cases could have died out without contributing to the main outbreak wave.

However, it can be argued that several imported cases may have contributed to the outbreak wave, and consequently may have accelerated the dynamic. This would imply that the outbreak would have started later.

As expected, the total number and intensity of imported cases responsible for the outbreak wave in France affects the starting date of the wave in the order of 4 to 9 days (fig. S6), which is similar to the sensitivity to the individual infectivity heterogeneity. Increasing the number of daily imported cases decreases the time to 100 detahs, even if the total number of imported cases is equivalent. Moreover, at a given number of daily imported cases, we can see that the difference between an importation during the first 5 days or during the first 30 days of the epidemic is very small. Therefore, only the importation of new infected individuals during the first days of the outbreak has an impact on the epidemic dynamic.

**Figure S6.**
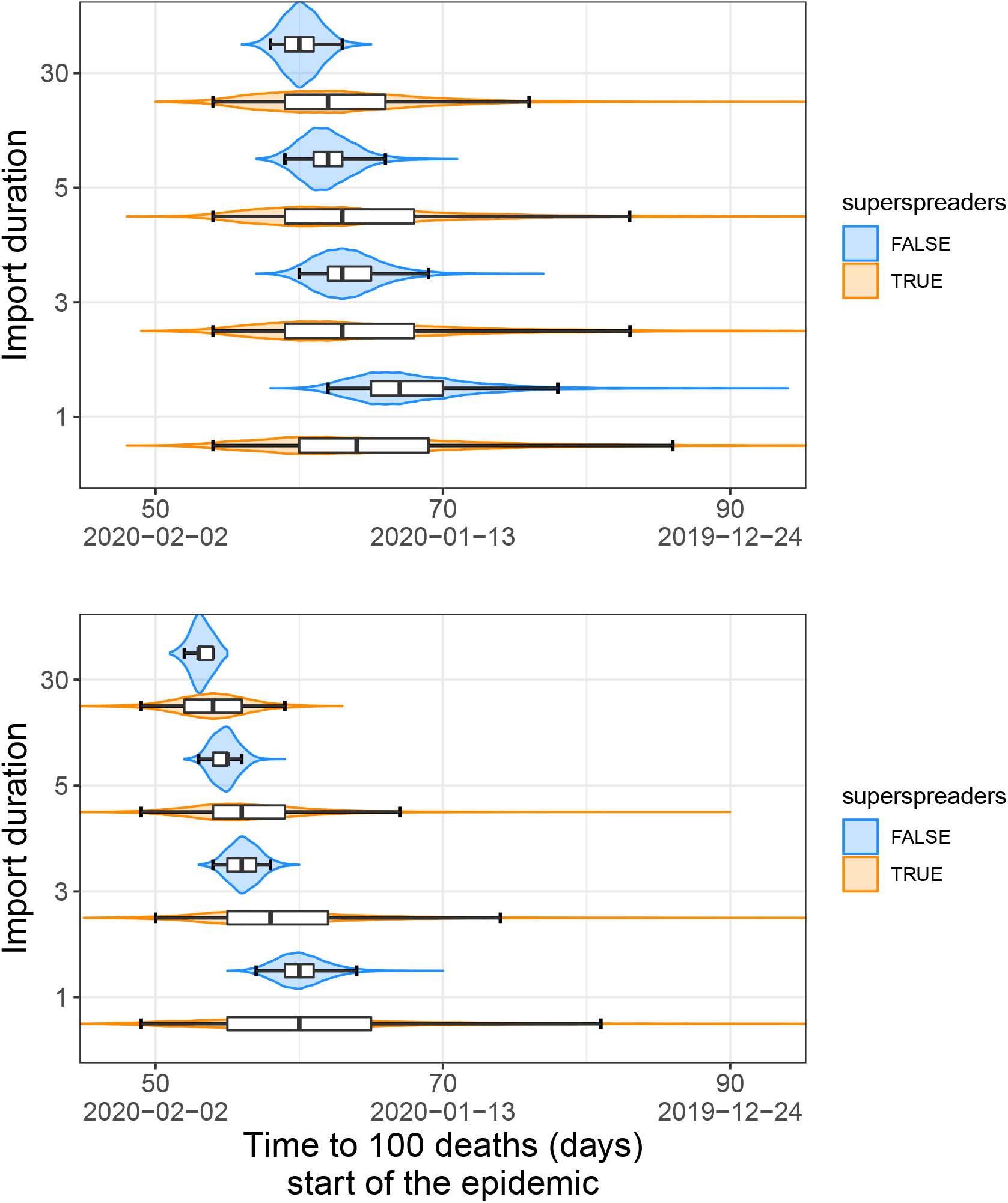
Time to 100 deaths distributions for various importation scenarios. Both panel display the effect of an increase in the period of first cases import on the distribution of the time to 100 deaths. In the upper panel, 1 cases was imported whereas in the lower panel 5 case were imported. The boxplots show the following quantiles: 2.5%, 25%, 50 %, 75%, 97.5 %.

#### Eradication and rebound risk with superspreading events

**Figure S7.**
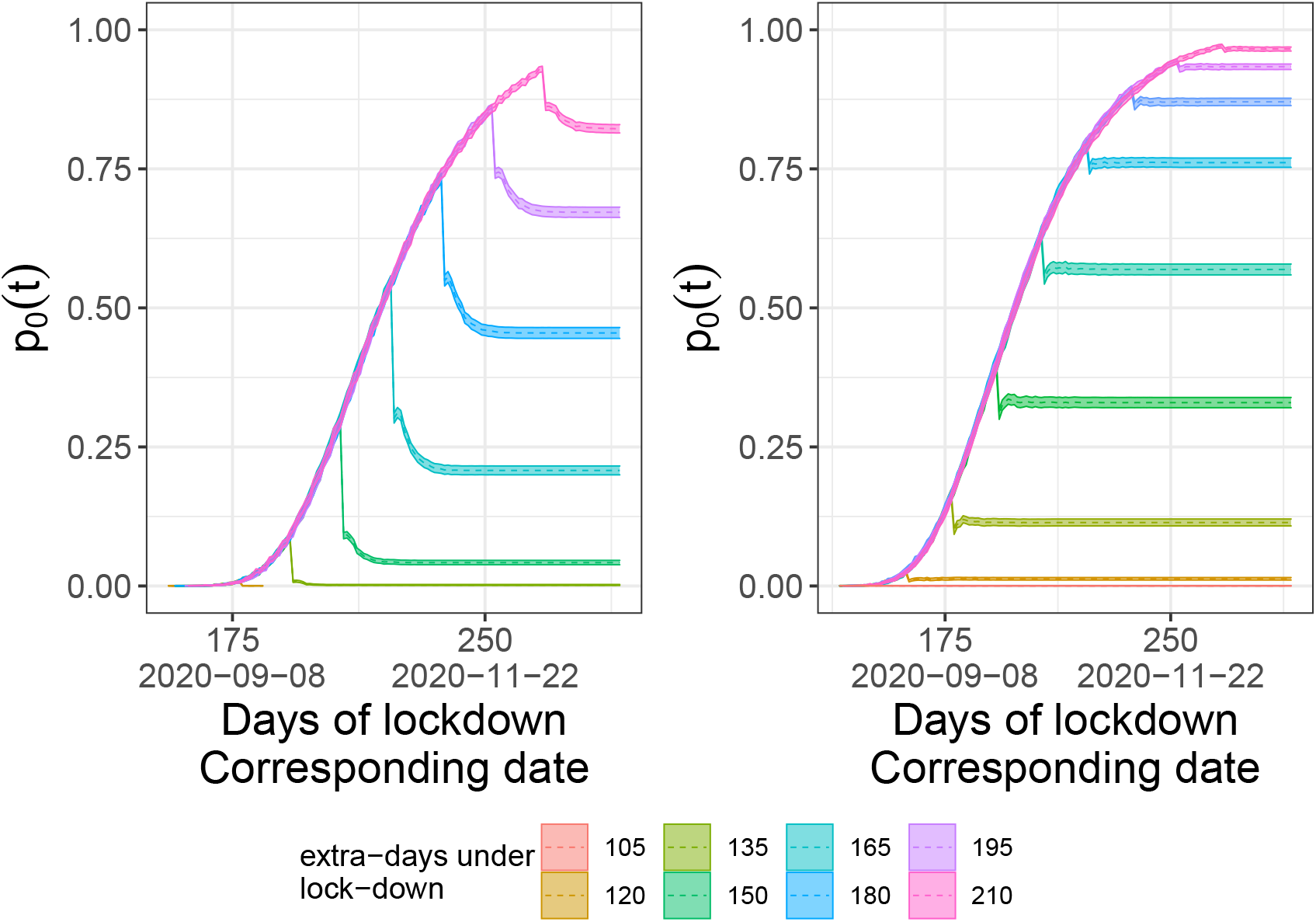
Variation of the estimated probability of having no new cases (*p*_0_(*t*)) with finite lock-down extension post 11 May: Here the x-axis corresponds to the number of days since the start of the lock-down (Mar 17) and to the corresponding date and the y-axis to estimation of the estimated probability of having no new cases. For the first 55 days we set the contact rate to its estimated value in France (*η*_*t*_ = 0.24), after this period we increase this rate to *η*_*>*55_ = 0.267 for a fixed duration (*d*) comprised between 15 days to 210 days. For the rest of the simulation we release all contact rate restrictions (*η*_*>*55 + *d*_ = 1). On the left panel we displayed the case without superspreading events and on the right panel when accounting for transmission heterogeneity. We plotted the estimation of the probability of epidemic extinction as dashed lines and the confidence interval as solid lines.

#### Eradication and lock-down initiation date

**Figure S8.**
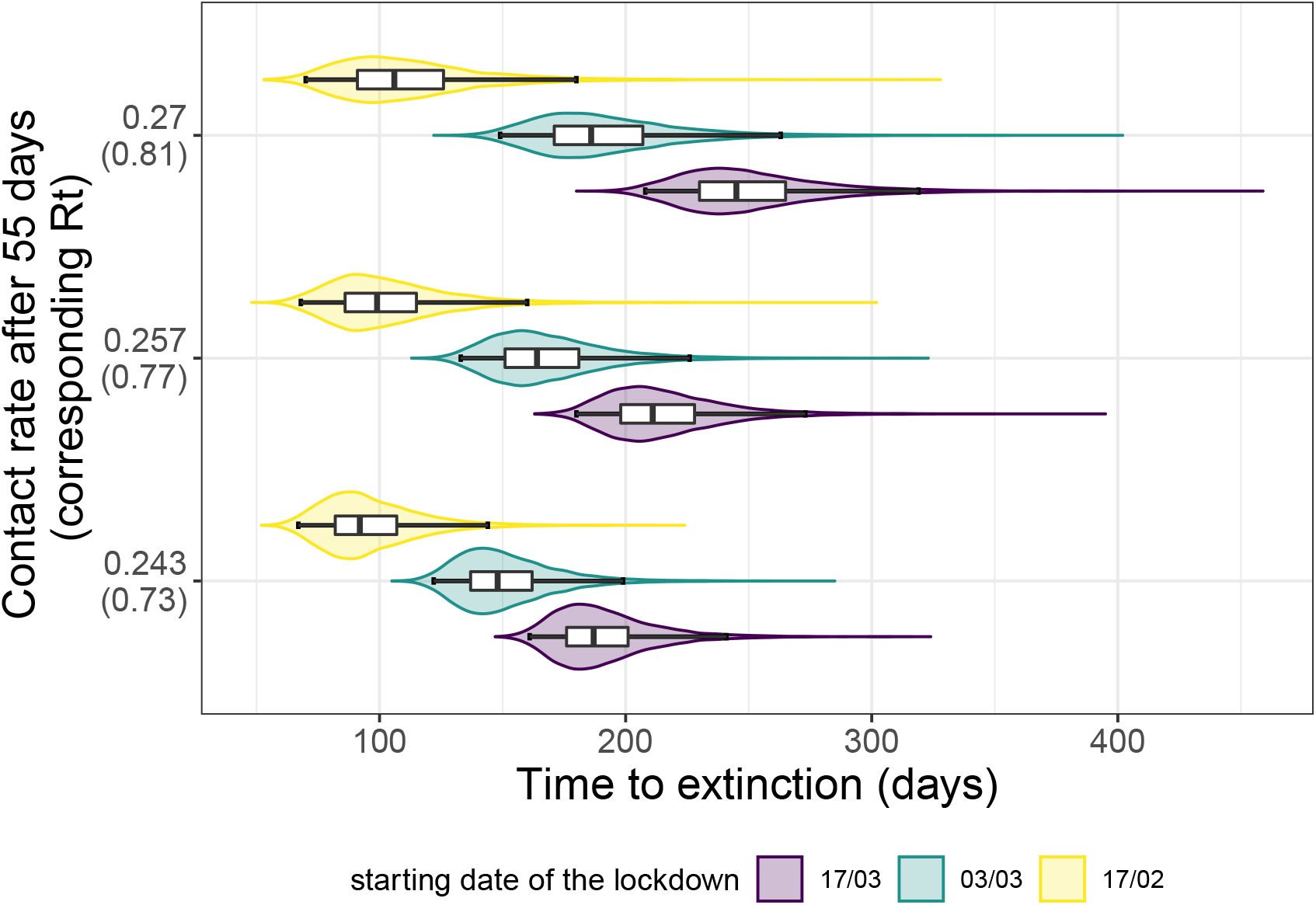
Effect of the lock-down intensity, stochasticity, and initiation date on the time to extinction (*τ*) without individual spreading heterogeneity assumption. The distributions of the time to extinction (in days since the start of the lock-down) for several contact rate restrictions post 55 first days are plotted on the Y-axis using violin plots and boxplots. In this graph we assume there is no individual spreading heterogeneity. The colors indicate the different initiation date of the lock-down: in purple it starts on Feb 17, green Mar 03 and yellow on Mar 17 (official start). The box extends from the lower to upper quartiles of the data. The whiskers expand from the 2.5% to the 97.5% quantiles.

#### Time to 20 new cases

**Figure S9.**
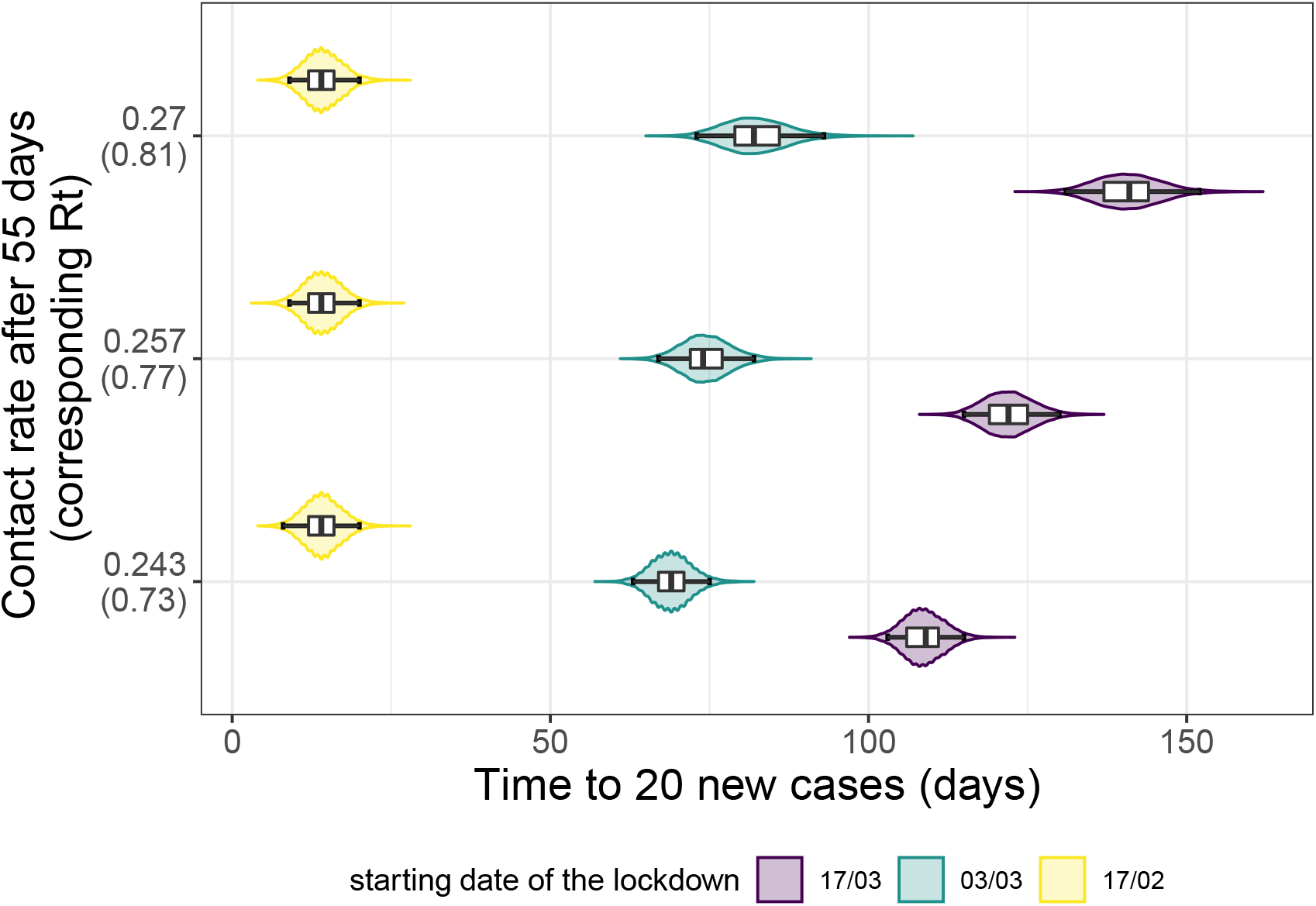
Effect of the lock-down intensity, stochasticity, and initiation date on the time to 20 new cases without superspreading events. The distributions of the time to 20 new cases (in days since the start of the lock-down) for several contact rate restrictions post 55 first days are plotted on the Y-axis using violin plots and boxplots. In this graph we assume no individual spreading heterogeneity. The colors indicate the different initiation date of the lock-down: in purple it starts on Feb 17, green Mar 03 and yellow on Mar 17 (official start). The box extends from the lower to upper quartiles of the data. The whiskers expand from the 2.5% to the 97.5% quantiles.

## References

Althouse BM, EA Wenger, JC Miller, SV Scarpino, A Allard, L Hébert-Dufresne, and H Hu (2020). Superspreading events in the transmission dynamics of SARS-CoV-2: Opportunities for interventions and control. PLOS Biology 18, e3000897. doi: 10.1371/journal.pbio.3000897.

Anderson RM and RM May (1991). Infectious Diseases of Humans. Dynamics and Control. Oxford: Oxford University Press.

Bedford T, AL Greninger, P Roychoudhury, LM Starita, M Famulare, ML Huang, A Nalla, G Pepper, A Reinhardt, H Xie, L Shrestha, TN Nguyen, A Adler, E Brandstetter, S Cho, D Giroux, PD Han, K Fay, CD Frazar, M Ilcisin, K Lacombe, J Lee, A Kiavand, M Richardson, TR Sibley, M Truong, CR Wolf, D. Nickerson, MJ Rieder, JA Englund, TSFS Investigators‡, J Hadfield, EB Hodcroft, J Huddleston, LH Moncla, N. Müller, RA Neher, X Deng, W Gu, S Federman, C Chiu, JS Duchin, R Gautom, G Melly, B Hiatt, P Dykema, S Lindquist, K Queen, Y Tao, A Uehara, S Tong, D MacCannell, GL Armstrong, GS Baird,HY Chu, J Shendure, and KR Jerome (2020). Cryptic transmission of SARS-CoV-2 in Washington state. Science. doi: 10.1126/science.abc0523.

Bénéteau T, B Elie, MT Sofonea, and S Alizon (Jan. 2021). Estimating Dates of Origin and End of COVID-19 Epidemics. en. medRxiv, 2021.01.19.21250080. doi: 10.1101/2021.01.19.21250080.

Britton T and G Scalia Tomba (Jan. 2019). Estimation in emerging epidemics: biases and remedies. Journal of The Royal Society Interface 16, 20180670. doi: 10.1098/rsif.2018.0670.

Danesh G, B Elie, Y Michalakis, MT Sofonea, A Bal, S Behillil, G Destras, D Boutolleau, S Burrel, AG Marcelin, JC Plantier, V Thibault, E Simon-Loriere, Sv der Werf, B Lina, L Josset, V Enouf, and S Alizon (2020). Early phylodynamics analysis of the COVID-19 epidemic in France. medRxiv 2020.06.03.20119925,. 3 peer-reviewed and recommended by PCI in Evolutionary Biology. doi: 10.24072/pci.evolbiol.100107.

Danesh G, E Saulnier, O Gascuel, M Choisy, and S Alizon (2020). Simulating trajectories and phylogenies from population dynamics models with TiPS. bioRxiv. doi: 10.1101/2020.11.09.373795.

Djaafara BA, N Imai, E Hamblion, B Impouma, CA Donnelly, and A Cori (2020). A Quantitative Framework to Define the End of an Outbreak: Application to Ebola Virus Disease. Am J Epidemiol, kwaa212. issn: 0002-9262, 1476-6256. doi: 10.1093/aje/kwaa212.

Eichner M and K Dietz (1996). Eradication of poliomyelitis: when can one be sure that polio virus transmission has been terminated? American journal of epidemiology 143, 816–822.

Endo A, Centre for the Mathematical Modelling of Infectious Diseases COVID-19 Working Group, S Abbott, AJ Kucharski, and S Funk (Apr. 2020). Estimating the overdispersion in COVID-19 transmission using outbreak sizes outside China. en. Wellcome Open Res 5, 67. issn: 2398-502X. doi: 10.12688/wellcomeopenres.15842.1.

Grant A (Apr. 2020). Dynamics of COVID-19 epidemics: SEIR models underestimate peak infection rates and overestimate epidemic duration. en. medRxiv. Publisher: Cold Spring Harbor Laboratory Press, 2020.04.02.20050674. issn: 2005-0674. doi: 10.1101/2020.04.02.20050674.

Hartfield M and S Alizon (2013). Introducing the outbreak threshold in epidemiology. PLoS Pathog. 6, e1003277. doi: 10.1371/journal.ppat.1003277.

He X, EHY Lau, P Wu, X Deng, J Wang, X Hao, YC Lau, JY Wong, Y Guan, X Tan, X Mo, Y Chen, B Liao, W Chen, F Hu, Q Zhang, M Zhong, Y Wu, L Zhao, F Zhang, BJ Cowling, F Li, and GM Leung (Apr. 2020). Temporal dynamics in viral shedding and transmissibility of COVID-19. en. Nat Med. Publisher: Nature Publishing Group, 1–4. issn: 1546-170X. doi: 10.1038/s41591-020-0869-5.

Hellewell J, S Abbott, A Gimma, NI Bosse, CI Jarvis, TW Russell, JD Munday, AJ Kucharski, WJ Edmunds, F Sun, S Flasche, BJ Quilty, N Davies, Y Liu, S Clifford, P Klepac, M Jit, C Diamond, H Gibbs, Kv Zandvoort, S Funk, and RM Eggo (Feb. 2020). Feasibility of controlling COVID-19 outbreaks by isolation of cases and contacts. English. The Lancet Global Health 0. issn: 2214-109X. doi: 10.1016/S2214-109X(20)30074-7.

Li Q, X Guan, P Wu, X Wang, L Zhou, Y Tong, R Ren, KSM Leung, EHY Lau, JY Wong, X Xing, N Xiang, Y Wu, C Li, Q Chen, D Li, T Liu, J Zhao, M Liu, W Tu, C Chen, L Jin, R Yang, Q Wang, S Zhou, R Wang, H Liu, Y Luo, Y Liu, G Shao, H Li, Z Tao, Y Yang, Z Deng, B Liu, Z Ma, Y Zhang, G Shi, TTY Lam, JT Wu, GF Gao, BJ Cowling, B Yang, GM Leung, and Z Feng (Jan. 2020). Early Transmission Dynamics in Wuhan, China, of Novel Coronavirus–Infected Pneumonia. en. New England Journal of Medicine. doi: 10.1056/NEJMoa2001316.

Linton NM, T Kobayashi, Y Yang, K Hayashi, AR Akhmetzhanov, Sm Jung, B Yuan, R Kinoshita, and H Nishiura (Feb. 2020). Incubation Period and Other Epidemiological Characteristics of 2019 Novel Coronavirus Infections with Right Truncation: A Statistical Analysis of Publicly Available Case Data. en. Journal of Clinical Medicine 9, 538. doi: 10.3390/jcm9020538.

Liu Y, RM Eggo, and AJ Kucharski (Mar. 2020). Secondary attack rate and superspreading events for SARS-CoV-2. English. The Lancet 395, e47. issn: 0140-6736, 1474-547X. doi: 10.1016/S0140-6736(20)30462-1.

Lloyd-Smith JO, SJ Schreiber, PE Kopp, and WM Getz (2005). Superspreading and the effect of individual variation on disease emergence. Nature 438, 355–9. doi: 10.1038/nature04153.

Max Roser Hannah Ritchie EOO and J Hasell (2020). Coronavirus Pandemic (COVID-19). Our World in Data. https://ourworldindata.org/coronavirus.

Nishiura H, NM Linton, and AR Akhmetzhanov (Mar. 2020a). Serial interval of novel coronavirus (COVID-19) infections. English. International Journal of Infectious Diseases 0. issn: 1201-9712. doi: 10.1016/j.ijid.2020.02.060.

Nishiura H, NM Linton, and AR Akhmetzhanov (Apr. 2020b). Serial interval of novel coronavirus (COVID-19) infections. International Journal of Infectious Diseases 93, 284–286. doi: 10.1016/j.ijid.2020.02.060.

Nishiura H, Y Miyamatsu, and K Mizumoto (Jan. 2016). Objective Determination of End of MERS Outbreak, South Korea, 2015. Emerging Infectious Diseases 22, 146–148. issn: 1080-6040. doi: 10.3201/eid2201.151383.

Plessis L du, JT McCrone, AE Zarebski, V Hill, C Ruis, B Gutierrez, J Raghwani, J Ashworth, R Colquhoun, TR Connor, NR Faria, B Jackson, NJ Loman, A O’Toole, SM Nicholls, KV Parag, E Scher, TI Vasylyeva, EM Volz, A Watts, II Bogoch, K Khan, C1GU(U Consortium†, DM Aanensen, MUG Kraemer, A Rambaut, and OG Pybus (Jan. 2021). Establishment and lineage dynamics of the SARS-CoV-2 epidemic in the UK. en. Science. Publisher: American Association for the Advancement of Science Section: Research Article. issn: 0036-8075, 1095-9203. doi: 10.1126/science.abf2946.

R Core Team (2020). R: A Language and Environment for Statistical Computing. R Foundation for Statistical Computing. Vienna, Austria.

Sofonea MT, B Reyné, B Elie, R Djidjou-Demasse, C Selinger, Y Michalakis, and S Alizon (2020). Epidemiological monitoring and control perspectives: application of a parsimonious modelling framework to the COVID-19 dynamics in France. medRxiv, 2020.05.22.20110593. doi: 10.1101/2020.05.22.20110593.

Sun K, W Wang, L Gao, Y Wang, K Luo, L Ren, Z Zhan, X Chen, S Zhao, Y Huang, Q Sun, Z Liu, M Litvinova, A Vespignani, M Ajelli, C Viboud, and H Yu (Nov. 2020). Transmission heterogeneities, kinetics, and controllability of SARS-CoV-2. en. Science. Publisher: American Association for the Advancement of Science Section: Research Article. issn: 0036-8075, 1095-9203. doi: 10.1126/science.abe2424.

Thompson RN, OW Morgan, and K Jalava (2019). Rigorous surveillance is necessary for high confidence in end-of-outbreak declarations for Ebola and other infectious diseases. Philosophical Transactions of the Royal Society B: Biological Sciences 374, 20180431. doi: 10.1098/rstb.2018.0431.

Trapman P, F Ball, JS Dhersin, VC Tran, J Wallinga, and T Britton (2016). Inferring R0 in emerging epidemics—the effect of common population structure is small. J. R. Soc. Interface 13, 20160288. doi: 10.1098/rsif.2016.0288.

Verity R, LC Okell, I Dorigatti, P Winskill, C Whittaker, N Imai, G Cuomo-Dannenburg, H Thompson, PGT Walker, H Fu, A Dighe, JT Griffin, M Baguelin, S Bhatia, A Boonyasiri, A Cori, Z Cucunubá, R FitzJohn, K Gaythorpe, W Green, A Hamlet, W Hinsley, D Laydon, G Nedjati-Gilani, S Riley, Sv Elsland, E Volz, H Wang, Y Wang, X Xi, CA Donnelly, AC Ghani, and NM Ferguson (Mar. 2020). Estimates of the severity of coronavirus disease 2019: a model-based analysis. English. The Lancet Infectious Diseases 0. issn: 1473-3099, 1474-4457. doi: 10.1016/S1473-3099(20)30243-7.

